# Symptom-based prediction model of SARS-CoV-2 infection developed from self-reported symptoms of SARS-CoV-2-infected individuals in an online survey

**DOI:** 10.1101/2020.11.25.20236752

**Authors:** Hansjörg Schulze, Daniel Hoffmann, Wibke Bayer

## Abstract

**Background:** Infections with the newly emerged severe acute respiratory syndrome virus 2 (SARS-CoV-2) have quickly reached pandemic proportions and are causing a global health crisis. First recognized for the induction of severe disease, the virus also causes asymptomatic infections or infections with mild symptoms that can resemble common colds. Since infections with mild course are probably a major contributor to the spread of SARS-CoV-2, better detection of such cases is important. To provide better understanding of these mild SARS-CoV-2 infections and to improve information for potentially infected individuals, we performed a detailed analysis of self-reported symptoms of SARS-CoV-2 positive and SARS-CoV-2 negative individuals.

**Methods:** In an online-based survey, 963 individuals provided information on symptoms associated with an acute respiratory infection, 336 of the participants had tested positive for SARS-CoV-2 infection, 107 had tested negative, and 520 had not been tested for SARS-CoV-2 infection.

**Results:** The symptoms reported most frequently by SARS-CoV-2 infected individuals were tiredness, loss of appetite, impairment of smell or taste and dry cough. The symptoms with the highest odds ratios between SARS-CoV-2 positive and negative individuals were loss of appetite and impairment of smell or taste. Based on the most distinguishing symptoms, we developed a Bayesian prediction model, which had a positive predictive value of 0.80 and a negative predictive value of 0.72 on the SARS-CoV-2 tested individuals. The model predicted 56 of 520 non-tested individuals to be SARS-CoV-2 positive with more than 75% probability, and another 84 to be SARS-CoV-2 positive with probability between 50% and 75%.

**Conclusions:** A combination of symptoms can provide a good estimate of the probability of SARS-CoV-2 infection.

## Background

The outbreak of severe acute respiratory syndrome coronavirus 2 (SARS-CoV-2), the virus associated with the coronavirus infectious disease 19 (COVID-19), at the end of 2019 [1] has quickly developed into pandemic proportions and was declared a public health emergency of international concern by the World Health Organization on January 30^th^ 2020 [2]. The first infections in Europe occurred in Italy and Germany in late January 2020 [3]. By March 2020, SARS-CoV-2 infection rose rapidly in many countries, and have started to do so again with the fall season in the northern hemisphere; as of November 6^th^ 2020, more than 48 million people have been infected by SARS-CoV-2, and more than 1.2 million SARS-CoV-2 related deaths have been reported [4].

Due to the rapid rise in demand for SARS-CoV-2 diagnostic tests and a resulting shortness of supplies, not all patients could be given access to SARS-CoV-2 testing in spring 2020, and a pre-screening was performed on the basis of symptoms, and prior residence in SARS-CoV-2 risk areas or prior contact to individuals who had tested SARS-CoV-2 positive. The lack of access to testing led to many uncertainties for patients with cold symptoms. From the first cases in China, the typical symptoms of SARS-CoV-2 infection were described as dry cough, fever, and pneumonia [5, 6]. These first reports of symptoms associated with SARS-CoV-2 infection / COVID-19 were based on symptoms found in hospitalized patients with severe disease. Notably, in the first reports on COVID-19, disease course was considered mild if patients did not require ventilation. It soon became obvious though that many SARS-CoV-2 infected individuals experienced asymptomatic or now commonly considered non-severe disease course that resembled common colds and did not require hospitalization [7]. Since the hospitalized and critically ill patients were of the greatest concern in the early days of the pandemic, the range of symptoms of SARS-CoV-2 infected individuals with mild symptoms was not the focus of attention. In following studies, it has been found that many SARS-CoV-2 infected patients experienced altered or completely lost sense of smell and / or taste [8, 9], which has since been regarded as a key indicator of SARS-CoV-2 infection.

To get a more detailed picture of the symptoms of SARS-CoV-2 infection, we created an online questionnaire and invited both tested and untested individuals to report their symptoms. The symptoms included in the survey comprised general symptoms such as fever, fatigue, headache and joint or muscle pains, and more specific eye, nose, throat, respiratory and gastrointestinal symptoms. The self-reported symptoms of SARS-CoV-2 positive- and negative-tested individuals were then used to fit a Bayesian model, which was used to predict probabilities of a SARS-CoV-2 infection of the non-tested survey participants.

## Methods

### Data collection

Data were collected in an online questionnaire based on LimeSurvey software hosted on the servers of the University of Duisburg-Essen. Participants of the online survey were recruited via public health offices of the city of Hamm (Northrhine-Westphalia, Germany), of the administrative district Soest (Northrhine-Westphalia, Germany), of the administrative district Hochsauerland (Northrhine-Westphalia, Germany) and the SARS-CoV-2 testing center in Lünen, administrative district Unna (Northrhine-Westphalia, Germany), as well as via social media. Participants were invited to complete the survey in case of a positive as well as negative SARS-CoV-2 test result, and also if they had not been tested but had cold symptoms. The data were collected between April 6^th^ and September 1^st^ 2020.

### Data analysis

Data were analyzed using R (version 3.6.3) and RStudio software and fsmb, plyr, splyr, tidyverse, randomForest, patchwork, rstanarm, ROCR, ggplot2, viridis and bayesplot packages. Only data from individuals who had completed the survey were included in the analysis.

For graphic representation of the survey results of tested individuals, data were sorted for the SARS-CoV-2 test result, the severity of the SARS-CoV-2 symptoms and the severity of fever using the tidyverse package, and a heatmap of the survey data was generated using ggplot2 package.

Frequencies of individual symptoms were calculated and visualized as heatmap using ggplot2 and viridis package. Odds ratios for individual symptoms were calculated as the conditional maximum likelihood estimate with 90% confidence interval using the pairwise.fisher.test command of R package fsmb, results were visualized using ggplot2.

Random Forest analysis of the survey data was performed using R package randomForest 4.6-14 [10] with default parameters. Importance of individual symptoms was characterized as mean decrease of Gini index on random permutations of case labels.

A Bayesian regression model was fitted with R package rstanarm version 2.21.1 [11] using the function stan_glm with default priors, on all or 10 selected symptoms of the tested individuals’ data as indicated in the results section, using a posterior sample size of 4000. To adjust for differences in the number of SARS-CoV-2 positive and negative tested participants, the dataset from negative participants (n = 107) was increased by bootstrapping to match the number of positive participants (n = 336). Bootstrapping was performed 4 times to exclude a bias of the results, with similar outcome. The R package bayesplot [12] was used to confirm the model validity by posterior predictive checks. Performance of the Bayesian and Random Forest based models as ROC curves (true positive rate and false positive rate) and area under the ROC curve were calculated with R package ROCR [13]. The numbers of true positives (TP), false positives (FP), true negatives (TN) and false negatives (FN) was determined using posterior predictions > 0.5 as positives and posterior predictions ≦ 0.5 as negative; positive predictive value (PPV) was calculated as PPV = TP / (TP + FP), negative predictive value (NPV) was calculated as NPV = TN / (TN + FN). The credibility intervals for PPV and NPV were calculated using the Voila online tool [14], which is based on a Bayesian classifier uncertainty estimate method [15].

For prediction of SARS-CoV-2 infection in untested individuals, the Bayesian model was applied to the dataset from the untested individuals and the most likely SARS-CoV-2 infection state was calculated for each of these individuals. For visualization of the prediction outcome, a heatmap was generated using ggplot2 after sorting the non-tested individuals by the predicted probability of SARS-CoV-2 infection as calculated for the Bayesian model.

## Results

To obtain an overview of the subjective symptoms reported by SARS-CoV-2 infected individuals, an online survey was performed. Invitations to the survey were published on social media, and SARS-CoV-2 tested individuals were also directly invited to participate when they received notification of their test results, or by subsequent invitation, by local public health offices. Participants included both tested individuals, as well as non-tested individuals, who were considered separately in the analysis of the survey. The characteristics of the tested study population are represented in Table 1. In total, responses of 443 participants who had been tested for SARS-CoV-2 infection were included in the survey, 336 (75.8 %) had tested positive for SARS-CoV-2 infection, while 107 (24.2 %) of those participants had tested negative. 293 (66.4 %) of the tested participants were recruited via the public health office, whereas 150 (33.6 %) tested participants were recruited from social media.

**Table 1.** Characteristics of study cohort.

|  | **SARS-CoV-2**<br>**positive**<br>**n (% of total)** | **SARS-CoV-2**<br>**negative**<br>**n (% of total)** | **untested**<br>**n (% of total)** |
| --- | --- | --- | --- |
| **number** | 336 | 107 | 520 |
| **age (years)** |  |  |  |
| 18 – 29 | 55 (16.4%) | 18 (16.8%) | 92 (17.7%) |
| 30 – 39 | 45 (13.4%) | 36 (33.6 %) | 178 (34.2%) |
| 40 – 49 | 61 (18.2%) | 26 (24.3%) | 126 (24.2%) |
| 50 – 59 | 118 (35.1%) | 19 (17.8%) | 88 (16.9%) |
| 60 – 69 | 41 (12.2%) | 7 (6.5%) | 29 (5.6%) |
| 70 – 79 | 9 (2.7%) | 1 (0.94%) | 5 (1.0%) |
| > 80 | 7 (2.1%) | 0 (0%) | 2 (0.4%) |
| **gender** |  |  |  |
| female | 189 (56.3%) | 81 (75.7%) | 409 (78.7%) |
| male | 147 (43.8%) | 25 (23.4%) | 109 (21.0%) |
| diverse | 0 (0%) | 1 (0.9%) | 2 (0.4%) |
| **smoking** | 13.1% | 23.4% | 30.2% |
| **hayfever\*** | 19.1% | 23.4% | 25.6% |

\* hayfever relevant at the time of infection

Of the 336 SARS-CoV-2 positive individuals, only 18 (5.3 %) reported no symptoms, whereas 141 (42.2 %) and 102 (30.1 %) reported their symptoms as mild or moderate, respectively, and 49 (14.7 %) reported their symptoms as severe. 20 (5.9 %) individuals were treated in the hospital due to the SARS-CoV-2 infection, and another 6 (1.8 %) required intensive care treatment.

The symptoms reported by the tested study population are shown as a heatmap in Figure 1, frequencies of the symptoms reported by SARS-CoV-2 positive and negative individuals are presented in Figure 2A. Clearly, a majority in both SARS-CoV-2 positive and negative individuals reported general symptoms such as tiredness and lethargy. The presence of eye symptoms in both groups was low, and also most nose symptoms were reported only by a minority of participants. Interestingly, about two thirds of SARS-CoV-2 positive individuals reported an impaired or lost sense of smell or taste. Dry cough was reported by almost 60% of both SARS-CoV-2 positive and negative individuals, whereas most SARS-CoV-2 positive individuals did not report other throat symptoms including throat pain. Approximately one third of individuals reported breathing problems, and also diarrhea was reported by approximately a third of SARS-CoV-2 positive and negative individuals.

**Figure 1.**
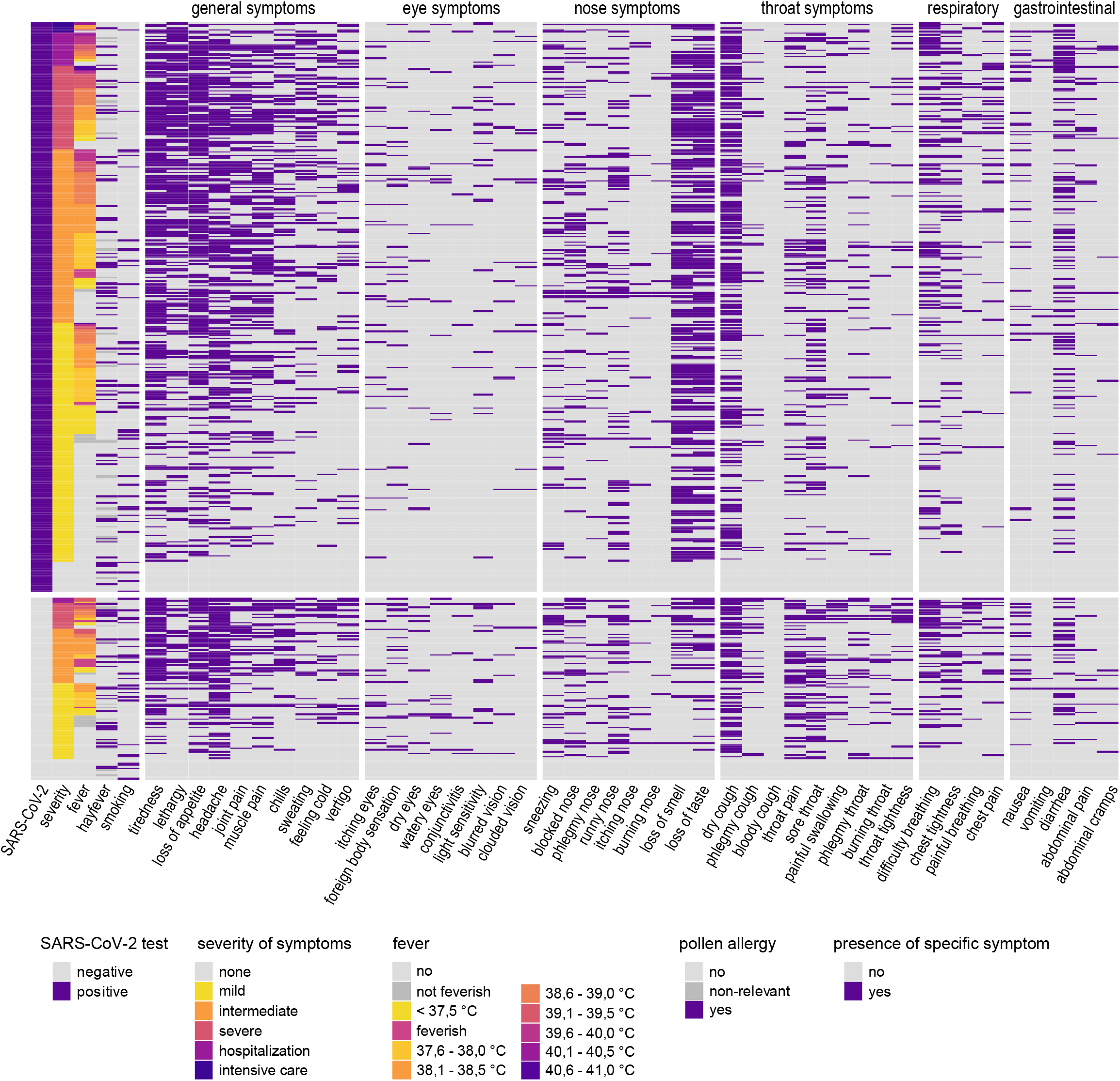
Heatmap of symptoms reported by SARS-CoV-2 positive and negative individuals. Symptoms reported by SARS-CoV-2 positive (top) and SARS-CoV-2 negative individuals (bottom). Subjects were sorted according to responses concerning severity and presence of fever. feverish/not feverish: subjective judgement, participant did not measure their body temperature.

**Figure 2.**
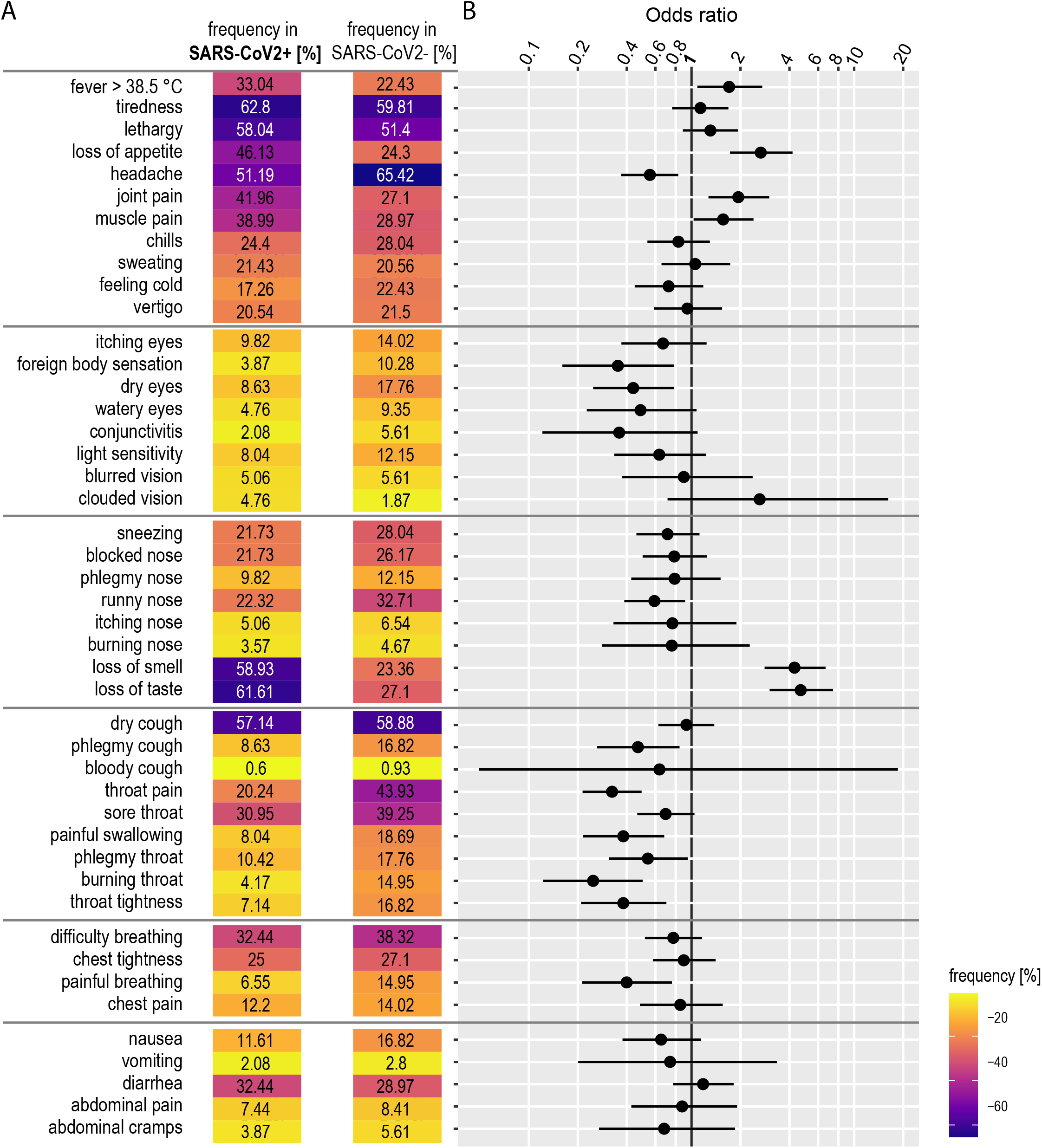
Frequencies and odds ratio of symptoms. (A) Frequencies for each symptom were calculated for SARS-CoV-2 positive and for SARS-CoV-2 negative individuals, respectively. (B) Odds ratios of individual symptoms and 90% confidence intervals.

The frequency of symptoms in SARS-CoV-2 positive and negative individuals were used to calculate the odds ratios for the individual symptoms (Figure 2B). Since the distribution of many symptoms was similar in SARS-CoV-2 positive and negative individuals, many odds ratios are close to 1. Clearly, the highest odds ratios were observed for impaired or lost sense of smell (estimate: 4.7; 90% confidence interval (CI): 2.8 – 8.1) and impaired or lost sense of taste (4.3; 90% CI: 2.6 – 7.2), whereas the other nose symptoms as well as eye and throat symptoms were found to have odds ratios below 1. The lowest odds ratios were observed for throat pain and burning sensation in the throat. The odds ratio of dry coughing is also slightly below 1 (0.9; 90% CI: 0.6 – 1.5), which may likely be an underestimation due to a bias in eligibility criteria at the time of testing.

For the development of a prediction model, we performed a Bayesian regression analysis of all symptoms and plotted them with their odds ratio in combination with the overall frequency (Figure 3A). As we have an imbalanced data set with less negative than positive tests, the resulting model would have been biased towards positive predictions. To avoid this, we fitted the model with two different approaches. In one, we increased the number of negatives by bootstrapping to match the number of positives, in the other we decreased the number of positives to match the number of negatives. Both approaches gave comparable results, but we favored the first one as it uses all the available data and is less prone to the introduction of a bias due to sample selection. For the final model, we selected symptoms that had marginal posterior probabilities of 90% or more to be either positively or negatively associated with positive or negative test results with mean posterior probabilities of > 0.5 or < -0.5 in the Bayes regression analysis, and were reported by at least 20% of the positive or negative individuals. We also performed a random forest regression analysis (Figure 3B), which confirmed the importance of the selected symptoms.

**Figure 3.**
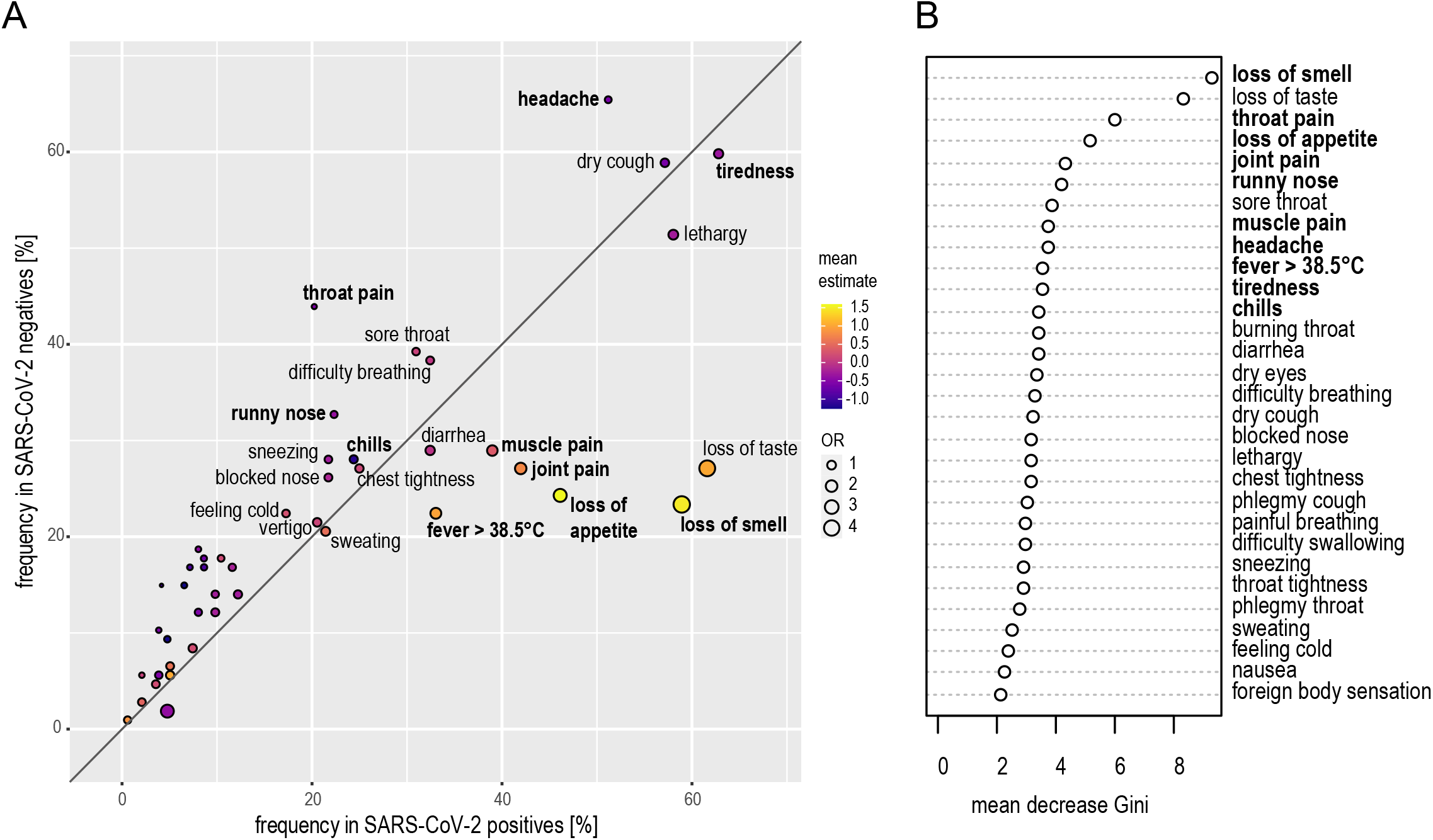
Bayesian regression and Random Forest-based selection of modelling factors. (A) For visual selection of symptoms for predictive modeling, the frequency of symptoms in SARS-CoV-2 negative individuals was plotted against the frequency in SARS-CoV-2 positive individuals for individual symptoms, odds ratio (OR) was visualized as dot size, Bayes model mean posterior probability estimate was visualized as color as indicated. Selected symptoms in bold letters. (B) Random Forest logistic regression analysis was performed to identify individual symptoms with importance for the development of a predictive model; the figure shows the mean decrease of the Gini index for the individual symptoms. Selected symptoms in bold letters.

Based on these analyses, we selected the ten symptoms impaired or lost sense of smell, throat pain, loss of appetite, joint pain, runny nose, muscle pain, headache, fever higher than 38.5 °C, tiredness and chills. As stated above, the high representation of dry cough in SARS-CoV-2 uninfected symptomatic individuals is likely due to the fact that it was communicated very early on as a typical symptom of SARS-CoV-2 infection, and was therefore a criterion for testing. Furthermore, we observed a strong correlation between impaired or lost sense of taste and sense of smell (Pearson’s r = 0.72). Therefore, neither dry cough nor loss of taste were included in the final Bayes model.

We used the selected ten symptoms to fit the final Bayesian regression model (Figure 4A). On the tested cohort data set, the model gave an area under the receiver operating characteristic curve of 0.80 (AUC; Figure 4B), a positive predictive value (PPV) of 0.80 (95% credible interval: 0.74 – 0.84) and a negative predictive value (NPV) of 0.72 (95% credible interval: 0.68 – 0.77). We applied the Bayesian model to the non-tested individuals who participated in the survey. The characteristics of this second study population are represented in Table 1. In total, responses of 520 participants who had not been tested for SARS-CoV-2 infection were included in the survey. Using the Bayesian model described above, we calculated that 56 (10.8%) were SARS-CoV-2 positive with a probability of more than 75% (Figure 4C). Another 84 individuals (16.2%) were predicted as SARS-CoV-2 positive with a probability between 50% and 75%. For the other 380 individuals (73.1%), the probability of SARS-CoV-2 infection based on the Bayesian model was less than 50%.

**Figure 4.**
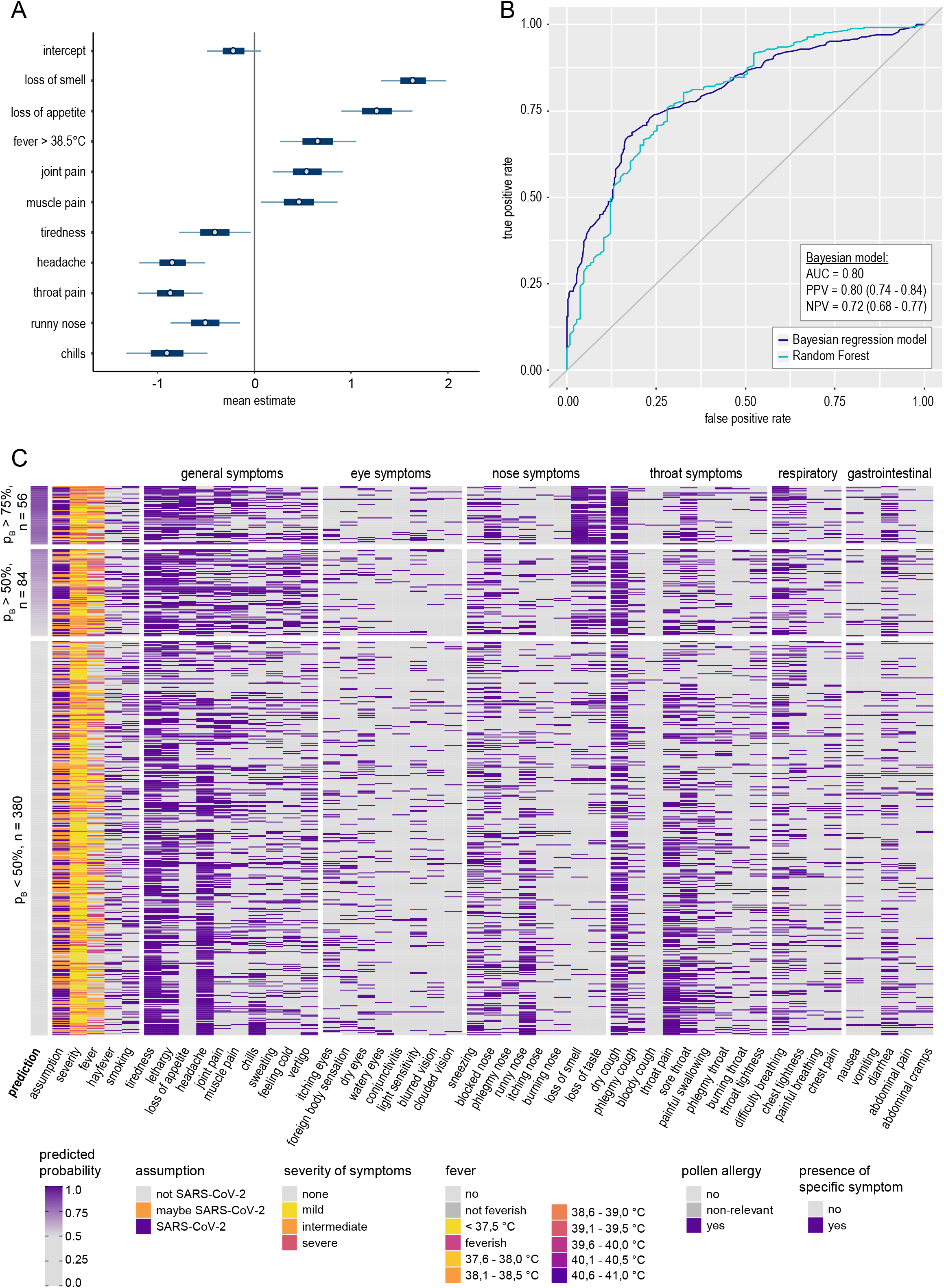
Heatmap of predicted probability of SARS-CoV-2 infection and of symptoms reported by non-tested individuals. (A) Bayesian regression analysis was performed using the indicated symptoms as priors, mean estimates and 90% highest posterior density intervals are shown. (B) Performance of the Bayesian model and the Random Forest analysis are shown as receiver operating characteristic curves; AUC (area under the curve), PPV (positive predictive value) and NPV (negative predictive value) with 95% credibility intervals of the Bayesian model on the model dataset of tested individuals are shown. (C) The dataset of non-tested individuals (n = 520) was submitted to the Bayesian model, and symptoms are shown for all individuals sorted from highest predicted probability (pB = Bayesian model prediction; top) to lowest predicted probability (bottom).

## Discussion

The data presented here provide a very detailed picture of the symptoms experienced by SARS-CoV-2 infected individuals. To our knowledge, we are providing the most detailed symptoms overview reported so far, having queried 45 symptoms ranging from general symptoms to specific eye, nose, throat, respiratory and gastrointestinal symptoms.

The data collected in this survey show that a range of symptoms is observed in a majority of patients, such as an impaired or lost sense of smell or taste, tiredness, lethargy, loss of appetite, joint or muscle pains and dry coughing. For some symptoms, we found surprisingly low frequencies in infected individuals and therefore low odds ratios, including all throat symptoms other than dry cough. Furthermore, only roughly one fifth of SARS-CoV-2 infected individuals experienced throat pain, which could be expected to be much higher since virus is routinely identified in throat swabs, showing that it is indeed infecting throat tissue.

For some symptoms, a bias may have been introduced into the dataset at two levels: some symptoms had been described early in the pandemic as typical for SARS-CoV-2 infection, and have therefore been criteria for testing, and on the other hand, the presence of these symptoms and the testing procedure itself may have increased the awareness of the affected individuals and increased their motivation to participate in this study. We expect that such a bias might exist for the symptoms “dry cough” and “difficulty breathing”. The odds ratios reported here may therefore be underestimates.

While many of the symptoms reported by SARS-CoV-2 positive individuals taken on their own are not useful for a prediction of SARS-CoV-2 infection, a prediction model based on 10 symptoms as we present here can provide a good estimate of the probability of a SARS-CoV-2 infection. The model therefore may provide useful guidance for personal behavior when symptoms are experienced as well as in the allocation of tests. It has to be noted though that while the positive predictive value of the model we describe here is rather high (0.80), the negative predictive value is lower (0.72). When used in the future for personal risk assessment, a predicted low probability of SARS-CoV-2 infection should therefore still be interpreted with caution and personal protection measures should be maintained.

Other studies have been performed investigating symptoms of SARS-CoV-2 and their predictive value. Studies in the early days of the COVID-19 pandemic were reporting symptoms of hospitalized patients and thus missed the symptoms that are associated with mild disease courses [5, 6, 16]. Since then, multiple other studies have been performed that focused on the symptoms experienced by SARS-CoV-2 infected individuals at a community level [8, 9, 17-21]. A large study has been performed using a mobile phone app that had more than seven thousand SARS-CoV-2 tested participants from the United Kingdom and the United States [19]. In this study, the authors found frequencies for the symptoms “loss of smell and taste”, “fever”, “skipped meals” and “diarrhea” that are comparable to the frequencies reported by us and reported positive odds ratios for these symptoms, with the highest odds ratio for “loss of smell and taste”. The authors report a similar frequency of “persistent cough” in the SARS-CoV-2 positive individuals as we found for “dry cough”; in contrast to our data, the frequency in SARS-CoV-2 negative individuals was lower than in our cohort, leading to a low positive odds ratio for the persistent cough, in contrast to our findings. The authors provided a prediction model based on age, sex, loss of smell and taste, persistent cough, severe fatigue and skipped meals, which obtained a positive predictive value of 0.69 and a negative predictive value of 0.75.

In another large study, only a small number of symptoms were surveyed: sore throat, cough, shortness of breath, loss of smell or taste, and fever [20]. Interestingly, the authors report distinctly lower frequencies for all these symptoms in their SARS-CoV-2 infected individuals, which were mostly only half of the frequencies we observed in our study and as low as only 2% of respondents reporting a fever of 38°C or higher, compared to more than 33% observed by us. This study also included a large number of individuals who were COVID-19 undiagnosed, who were ten times the number of diagnosed individuals, however more than 95% of the undiagnosed individuals reported to feel well and not have any of the queried symptoms, creating an imbalance between the SARS-CoV-2 positive and negative study population. The model proposed in that study that relies on this small number of symptoms would likely prove less effective when applied to a cohort comprising more symptomatic individuals.

In contrast to these two large-scale studies, we did not include age and sex in our models but included more symptoms, which resulted in a higher positive predictive value. Some of the symptoms included in our model are more subjective and may vary in strength, for example tiredness and loss of appetite. However, we think that modeling a prediction on this higher number of symptoms provides a more precise model that can be used for a first prediction of the probability of SARS-CoV-2 infection, guiding decisions for self-isolation and testing.

## Conclusions

Our data shows that many symptoms reported by SARS-CoV-2 infected individuals are rather unspecific and on their own are not amenable to a clear distinction of SARS-CoV-2 infection from infections with other viruses causing common cold symptoms. Nevertheless, our Bayesian model based on 10 symptoms provides a good prediction of probability of SARS-CoV-2 infection. The model can provide a probability estimate and provide guidance in test allocation where testing capacities are limited.

## Declarations

### Ethics approval and consent to participate

The study was approved by the local Ethics Committee of the Medical Faculty of the University Duisburg-Essen (approval number 20-9233-BO)

### Consent for publication

Not applicable

### Availability of data and materials

The datasets used and/or analyzed during the current study are available from the corresponding author on reasonable request.

### Authors’ contributions

HS recruited participants, analyzed data and contributed to writing the manuscript. DH analyzed data and contributed to writing the manuscript. WB conceived of the study, created the online survey, analyzed data and wrote the manuscript.

## Data Availability

The datasets used and/or analyzed during the current study are available from the corresponding author on reasonable request after publication.

## Acknowledgements

The authors thank all study participants for taking the time to complete the online questionnaire and contributing to our advancement in the recognition of SARS-CoV-2 symptoms. The authors also thank the employees of the public health offices in Hamm (Germany), Soest (Germany), Hochsauerlandkreis (Germany) and Dr. Hüning and nurses of the SARS-CoV-2 treatment center in Lünen (Germany) for distributing invitations to the online survey.

